# Efficacy and Safety of Oral Gene III^®^ L-Ergothioneine Capsules in Primary Dysmenorrhea: A Randomized, Double-Blind, Placebo-Controlled Clinical Trial

**DOI:** 10.64898/2026.03.26.26349375

**Authors:** Cong Guo, Wei Liu, Wei Ding, Juan Cao, Tong Tong, Fengjuan Liu, Guohua Xiao

**Author notes:** Corresponding author: Guohua Xiao.

## Abstract

**Purpose:** To evaluate the efficacy and safety of oral L-ergothioneine (EGT) in alleviating pain and associated symptoms in women with primary dysmenorrhea (PD).

**Methods:** In this randomized, double-blind, placebo-controlled trial, 40 women with PD (aged 18–30 years) were randomized (1:1) to receive EGT capsules (120 mg/day) or a matching placebo for 3 consecutive menstrual cycles. Outcomes evaluated at baseline and post-cycle included peak pain (Visual Analog Scale, VAS), Dysmenorrhea Symptom Score, and the COX Menstrual Symptom Scale (CMSS).

**Results:** EGT significantly improved PD symptoms over 3 cycles. Mean VAS for peak pain decreased from 4.80 ± 1.12 to 2.32 ± 1.59 in the EGT group (p < 0.001), compared to a non-significant reduction (4.10 ± 1.30 to 3.45 ± 1.69) in the placebo group. The between-group difference at cycle 3 was significant (p < 0.01). A linear mixed-model confirmed a significant Time × Group interaction (p < 0.001), showing an accelerated decline in symptom severity for EGT. Furthermore, 84% of EGT-treated patients achieved ≥50% VAS reduction versus 35% in the placebo group (p = 0.003). Serum inflammatory biomarkers showed no significant between-group differences or correlation with VAS improvements, suggesting EGT’s analgesic effects likely operate via cytoprotective pathways independent of classical inflammatory cascades. No adverse events were reported.

**Conclusion:** Oral EGT supplementation (120 mg/day) effectively and progressively mitigates menstrual pain and systemic symptoms in PD, offering a well-tolerated, non-pharmacological nutritional intervention.

**Trial Registration:** ChiCTR2500112557; Retrospectively registered on 2025-11-17.

## 1. Introduction

Primary dysmenorrhea (PD) is the most common gynecological disorder among women of reproductive age, characterized by cramping lower abdominal pain occurring just before or during menstruation, in the absence of identifiable pelvic pathology [1, 2]. Epidemiological studies indicate that the global prevalence of PD ranges from 16% to over 90%, significantly impairing patients’ quality of life, academic performance, and work productivity [3, 4]. The core pathogenesis of PD is driven by the overproduction of endometrial prostaglandins (particularly PGF2α and PGE2), which induces uterine hypercontractility, local ischemia, and hypoxia [5]. Accumulating evidence reveals that these ischemic events trigger an inflammatory cascade and severe oxidative stress. Elevated levels of reactive oxygen species (ROS) and lipid peroxidation markers, alongside depleted antioxidant reserves, are closely linked to the severity of dysmenorrheic pain [6, 7].

Given the limitations and adverse effects of chronic NSAID use, an increasing number of women are turning to nutritional strategies and dietary supplements to manage menstrual discomfort safely [8]. Among these functional dietary components, L-ergothioneine (EGT) has emerged as a unique naturally occurring, highly stable, sulfur-containing amino acid derivative synthesized primarily by fungi and mycobacteria [9]. EGT represents a novel dietary micronutrient that cannot be synthesized by the human body and must be acquired strictly through diet or targeted nutritional supplementation [9]. Unlike conventional dietary antioxidants, EGT exhibits a unique cellular uptake mechanism mediated by a highly specific organic cation transporter, OCTN1 (encoded by the SLC22A4 gene) [10]. Because OCTN1 is heavily expressed in tissues subjected to high oxidative stress, EGT actively accumulates within mitochondria and cells, where it scavenges free radicals, protects DNA and proteins from oxidative damage, and helps maintain a balanced inflammatory response [11, 12]. Despite these promising cytoprotective properties, clinical studies evaluating EGT as a targeted nutritional strategy for menstrual discomfort remain absent. Therefore, this clinical trial was designed to investigate the efficacy and safety of oral EGT capsules in supporting uterine comfort and mitigating PD symptoms.

While the specific expression of the OCTN1 transporter in human myometrium requires further mapping, existing literature confirms its ubiquitous presence in tissues susceptible to oxidative damage. Given that primary dysmenorrhea is characterized by acute oxidative stress and inflammation driving localized ischemia, we hypothesized that the targeted mitochondrial accumulation of EGT could mitigate these upstream triggers, thereby safely alleviating pelvic pain.

## 2. Materials and Methods

### 2.1 Study Design

This prospective, single-center, randomized, double-blind, parallel, placebo-controlled trial was conducted at the Qingdao Central Hospital of Rehabilitation University between July 2025 and December 2025. The protocol was approved by the hospital’s Clinical Trial Ethics Committee, and the study adhered strictly to the Declaration of Helsinki and Good Clinical Practice (GCP) guidelines. Written informed consent was obtained from all participants prior to screening.

### 2.2 Participants

A total of 40 women were enrolled. Inclusion criteria required participants to be nulliparous females aged 18 to 30 years, diagnosed with primary dysmenorrhea. Diagnosis was based on pain onset 1–2 years post-menarche, beginning just before or during menstruation, with no abnormal pelvic pathology confirmed by gynecological examination. Participants were required to have regular menstrual cycles (28 ± 7 days) and a baseline peak pain Visual Analog Scale (VAS) score of ≥ 40 mm based on the recall of the last three cycles. Exclusion criteria included secondary dysmenorrhea (e.g., endometriosis, adenomyosis, pelvic inflammatory disease); severe cardiovascular, hepatic, renal, or psychiatric disorders; pregnancy or lactation; alcohol or drug abuse; and the use of NSAIDs, traditional Chinese medicine, or other dysmenorrhea treatments within the past month. Participants were also excluded if they presented clinically significant abnormal baseline laboratory parameters (e.g., hemoglobin < 80 g/L, uncontrolled blood pressure). To minimize confounding factors, all participants were instructed to avoid strenuous exercise throughout the trial period. Additionally, the use of any concurrent treatments, including other dietary supplements, analgesics, or traditional Chinese medicines that might affect efficacy or safety evaluations, was strictly prohibited.

### 2.3 Interventions

Participants were randomly assigned (1:1 ratio) into two groups using block randomization: Experimental Group (n = 20): Received Gene III^®^ L-Ergothioneine Capsules (30 mg EGT per capsule). Participants took 2 capsules in the morning and 2 in the evening (total daily dose: 120 mg). The capsules were manufactured by Alpha Plus Tech, LLC, and provided by Jiangsu Gene III Biotechnology Co., Ltd. Placebo Group (n = 20): Received identically matched placebo capsules (0.3 g/capsule) following the exact dosing schedule. The placebo was manufactured by Lujianyuan (Xinxiang) Bioengineering Co., Ltd., and provided by Jiangsu Gene III Biotechnology Co., Ltd. Both EGT and placebo capsules were identical in appearance, packaging, and labeling to maintain the double-blind design. The intervention lasted for 3 consecutive menstrual cycles.

Supplementation compliance was strictly monitored throughout the trial. Participants were required to record their daily capsule intake in designated diary cards. At each follow-up visit,unused capsules and empty packaging were collected and counted by the investigators. Overall compliance was calculated as the percentage of capsules actually taken relative to the prescribed dose. In this study, the overall compliance rate exceeded 95% in both groups, indicating excellent adherence to the intervention protocol.

### 2.4 Outcome Measures

Clinical evaluations were conducted at baseline and at the end of the 1st, 2nd, and 3rd menstrual cycles. Visual Analog Scale (VAS): Evaluated peak pain intensity during each cycle. Dysmenorrhea Symptom Score: Graded the severity of core symptoms (abdominal pain, systemic symptoms, duration, and impact on daily life). COX Menstrual Symptom Scale (CMSS): Quantified the frequency and severity of dysmenorrhea-associated symptoms (e.g., nausea, fatigue, mood fluctuations). Exploratory Outcomes: Assessed at baseline and the third cycle, including inflammatory factors (TNF-α, IL-6, IL-1β, and Prostaglandin F2α) and breast ultrasound evaluations for nodule quantity and size. The detailed results of these exploratory mechanistic biomarkers will be reported in a separate manuscript.

### 2.5 Safety Assessment

Safety was evaluated using the Common Terminology Criteria for Adverse Events (CTCAE v5.0). Comprehensive physical examinations, vital signs, and laboratory tests were conducted at screening and at study exit (post-cycle 3).

### 2.6 Statistical Methods

Given the scarcity of previous clinical trials utilizing oral L-ergothioneine for primary dysmenorrhea, this study was designed as an exploratory pilot trial. A sample size of 40 participants (20 per group) was determined based on established methodological recommendations for pilot clinical trials, which suggest that 15–20 participants per group is adequate to evaluate initial clinical efficacy, feasibility, and safety trends before powering a larger, definitive multicenter trial.

Statistical analyses were performed using Python (SciPy, statsmodels) and SAS 9.4. Continuous variables were expressed as mean ± standard deviation (SD) or mean ± standard error of the mean (SEM). Within-group comparisons were conducted using the Wilcoxon signed-rank test or paired t-test based on the Shapiro–Wilk normality test. Between-group comparisons used the Mann–Whitney U test. Fisher’s exact test was used for categorical responder outcomes. Longitudinal VAS trajectories were modeled using a linear mixed model (LMM) with visit (0–3), group (EGT vs. placebo), and their interaction as fixed effects, and a random intercept per subject. The Time × Group interaction term served as the primary inferential test for differential intervention trajectories. Correlations between baseline VAS and intervention response were assessed by Spearman’s rank correlation (ρ_s_). A two-sided significance level of α = 0.05 was applied.

The sample size (n=40) was determined based on the exploratory nature of this pilot study. The LMM utilized a full analysis set (FAS) to account for all available data points, effectively handling the single missing observation at Cycle 3 for subject K007.

## 3. Results

### 3.1 Participant Flow and Baseline Characteristics

All 40 enrolled participants (20 per group) completed the 3-month intervention and follow-up phase (Figure 1). No participants dropped out. However, one subject in the EGT group (K007) missed the final assessment at Cycle 3. At baseline, there were no statistically significant differences between the groups regarding age, Body Mass Index (BMI), baseline pain intensity, or dysmenorrhea symptom scores, indicating high comparability prior to intervention.

**Figure 1.**
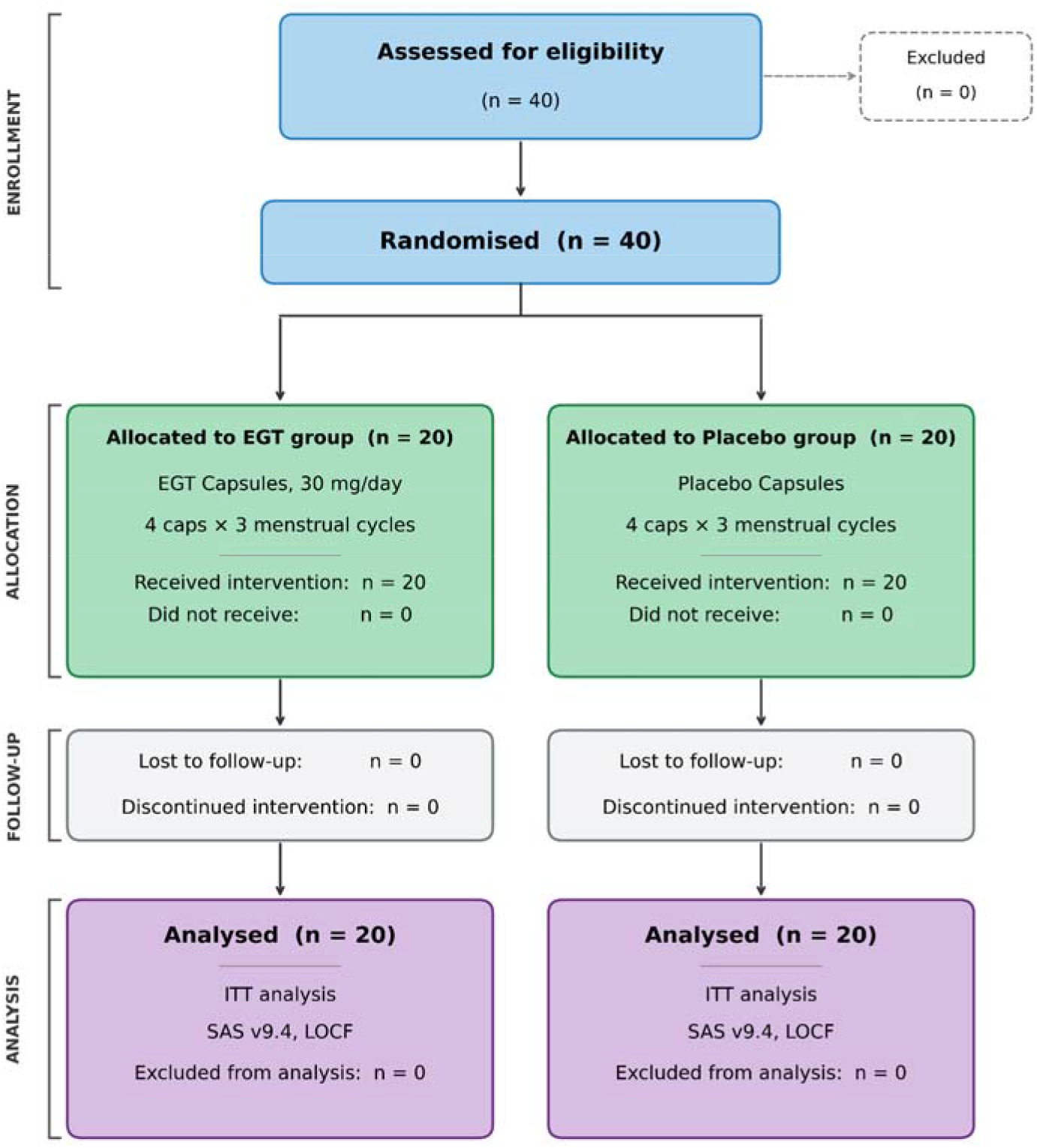
CONSORT flow diagram of the study. The diagram illustrates the progression of participants through the trial, including enrollment, randomization, allocation, follow-up, and data analysis.

**Table 1.**
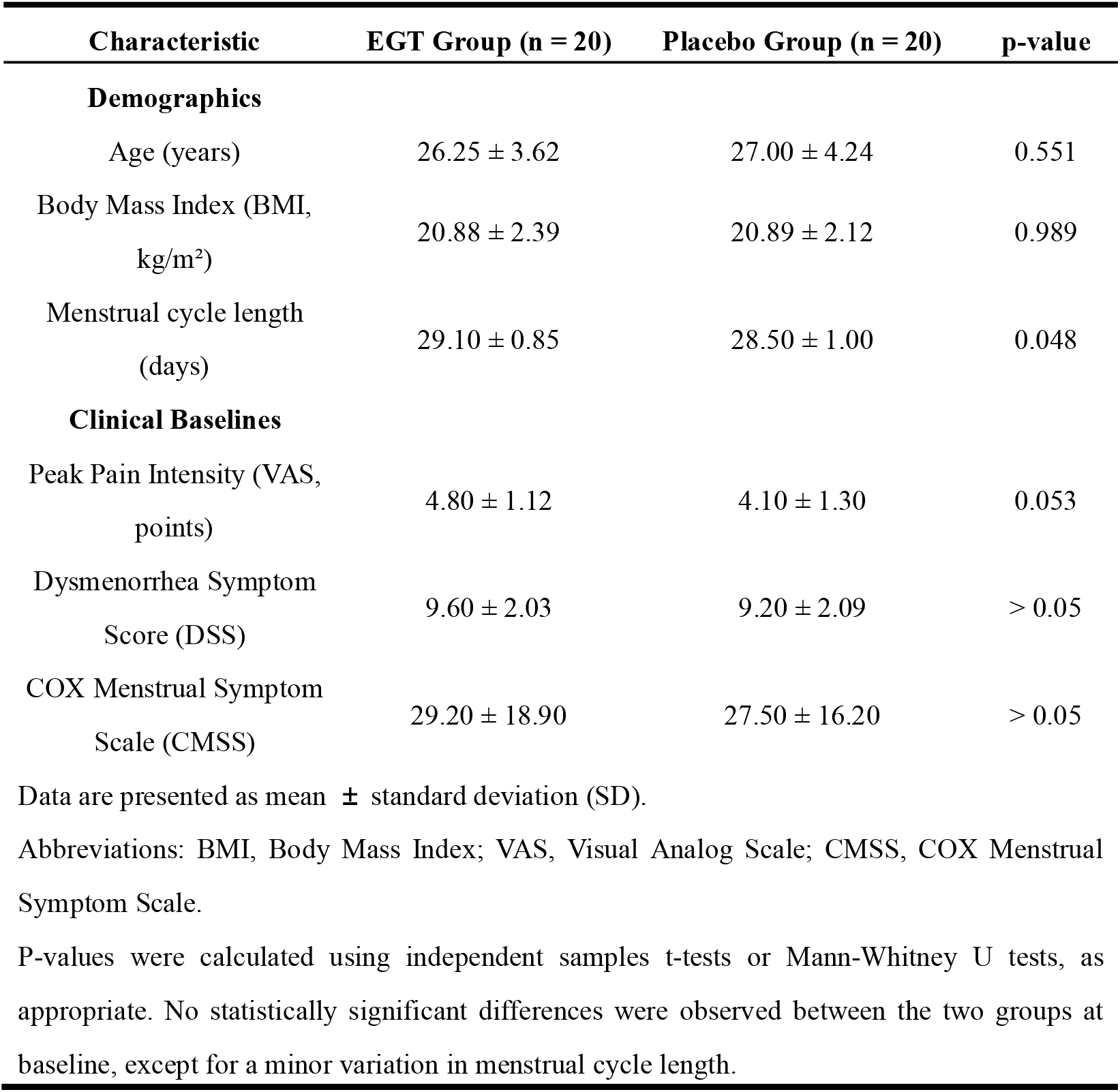
Baseline demographic and clinical characteristics of the study participants.

| Characteristic | EGT Group (n = 20) | Placebo Group (n = 20) | p-value |
| --- | --- | --- | --- |
| **Demographics** |  |  |  |
| Age (years) | 26.25 ± 3.62 | 27.00 ± 4.24 | 0.551 |
| Body Mass Index (BMI, kg/m<sup>2</sup>) | 20.88 ± 2.39 | 20.89 ± 2.12 | 0.989 |
| Menstrual cycle length (days) | 29.10 ± 0.85 | 28.50 ± 1.00 | 0.048 |
| **Clinical Baselines** |  |  |  |
| Peak Pain Intensity (VAS, points) | 4.80 ± 1.12 | 4.10 ± 1.30 | 0.053 |
| Dysmenorrhea Symptom Score (DSS) | 9.60 ± 2.03 | 9.20 ± 2.09 | > 0.05 |
| COX Menstrual Symptom Scale (CMSS) | 29.20 ± 18.90 | 27.50 ± 16.20 | > 0.05 |

Data are presented as mean ± standard deviation (SD).
Abbreviations: BMI, Body Mass Index; VAS, Visual Analog Scale; CMSS, COX Menstrual Symptom Scale.
P-values were calculated using independent samples t-tests or Mann-Whitney U tests, as appropriate. No statistically significant differences were observed between the two groups at baseline, except for a minor variation in menstrual cycle length.

### 3.2 Primary Efficacy Outcome: Peak Pain Intensity (VAS)

Over the 3-cycle intervention, the EGT group exhibited a progressive and statistically significant reduction in peak menstrual pain. The mean VAS score in the EGT group decreased from 4.8 ± 1.12 at baseline to 4.1 ± 0.99 at cycle 1, 3.6 ± 1.32 at cycle 2, and 2.3 ± 1.59 at cycle 3 (all P < 0.001 vs. baseline). Conversely, the placebo group demonstrated no significant intra-group change throughout the trial. By the end of the third cycle, the inter-group difference in VAS scores reached statistical significance (P = 0.0052), confirming the distinct analgesic efficacy of EGT over placebo. (Figure 2)

**Figure 2.**
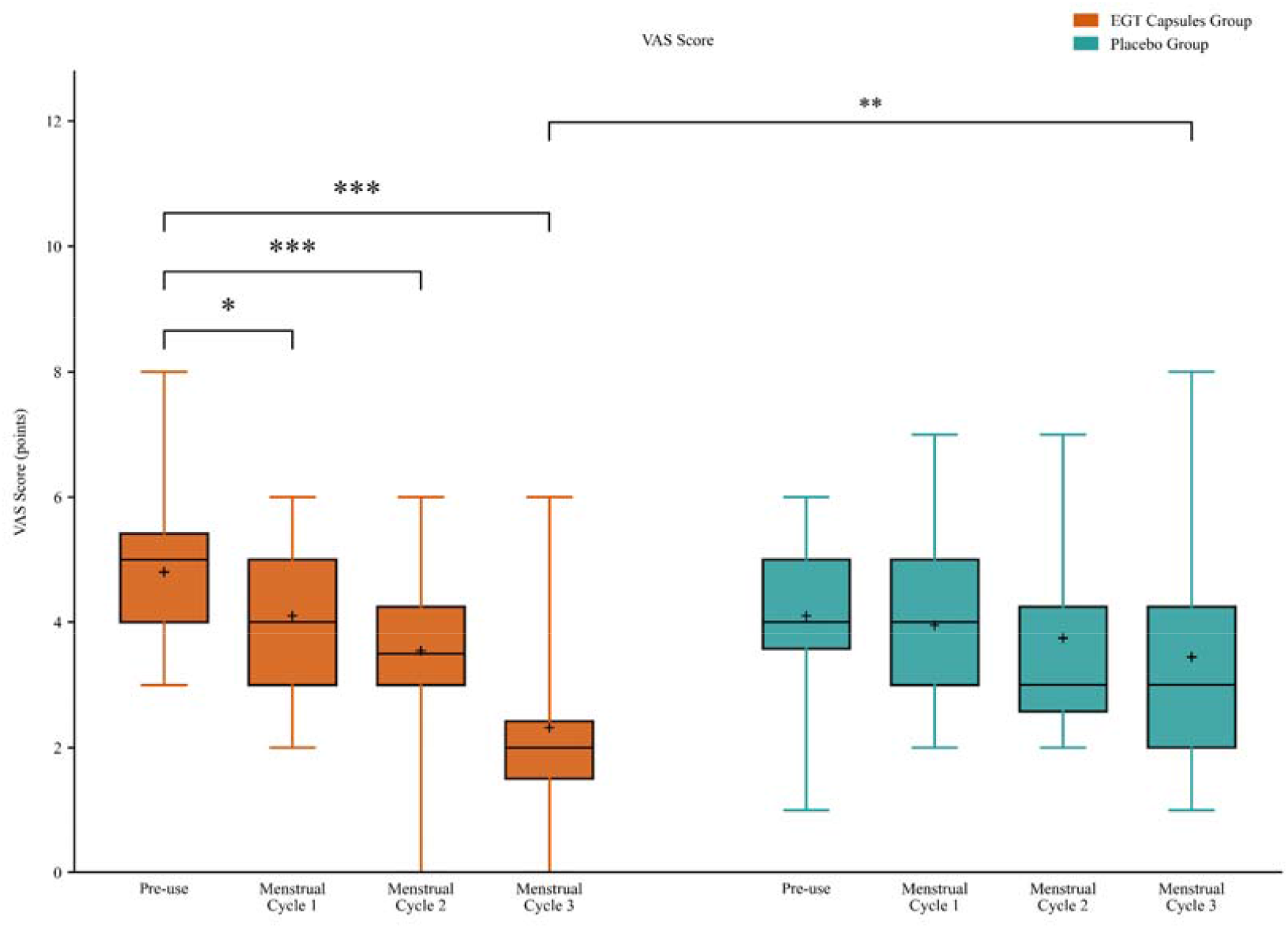
Effects of oral L-ergothioneine (EGT) on peak menstrual pain intensity. Data are presented as mean Visual Analog Scale (VAS) scores ± standard deviation (SD) at baseline (V0) and after Menstrual Cycles 1–3 for the EGT (blue circles, n = 20) and Placebo (orange squares, n = 20) groups. *p < 0.05, **p < 0.01, ***p < 0.001 indicate within-group significance compared to baseline. †p = 0.005 indicates between-group significance at Cycle 3.

### 3.3 Secondary Efficacy Outcomes: Dysmenorrhea Symptom Score and CMSS

Dysmenorrhea Symptom Score: The mean score consistently decreased from 9.6 ± 2.03 at baseline to 8.5 ± 2.56, 7.6 ± 2.66, and 5.7 ± 2.59 after cycles 1, 2, and 3, respectively (all P < 0.05 vs. baseline). Although the placebo group registered a slight, non-significant reduction, the intervention effect of EGT was significantly superior to the placebo at the final endpoint (P = 0.0282) (Figure 3A). CMSS: Systemic menstrual complaints were significantly mitigated in the EGT group. Total CMSS scores fell from 29.2 ± 18.9 at baseline to 16.4 ± 11.53 at cycle 3 (P < vs. baseline). No significant fluctuations in CMSS scores were observed in the placebo group. (Figure 3B)

**Figure 3.**
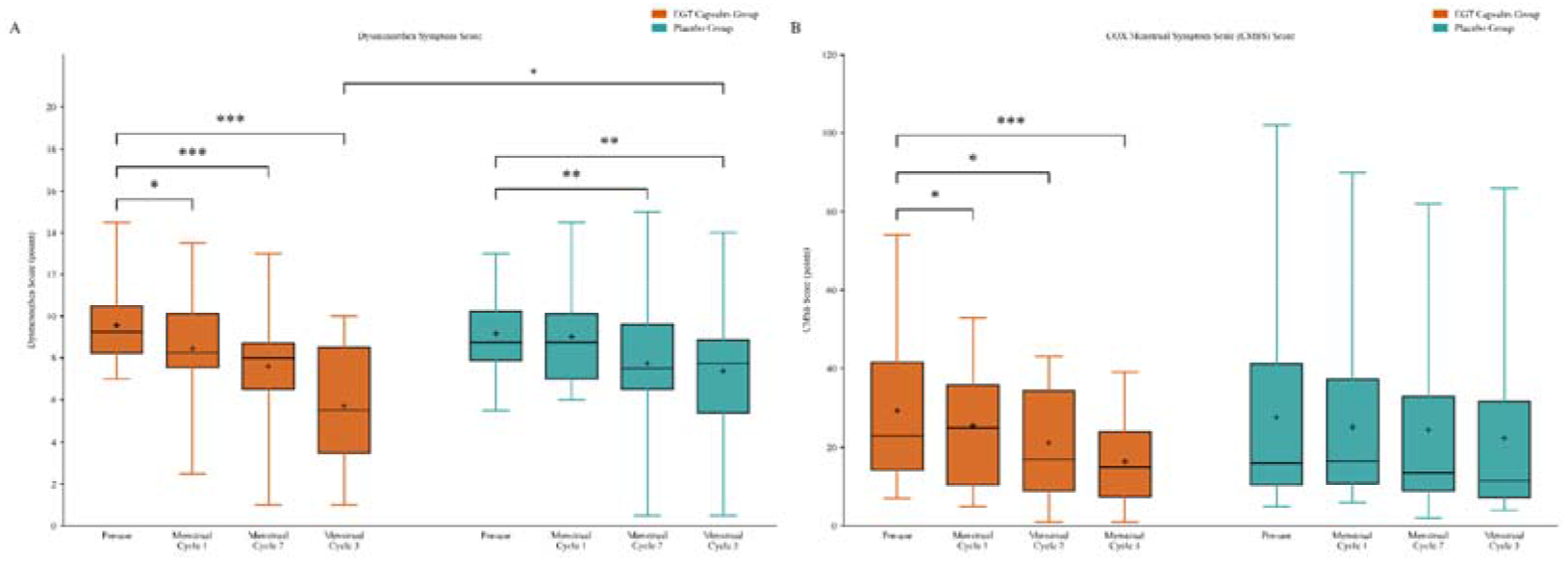
Effects of oral L-ergothioneine (EGT) on secondary efficacy outcomes. **(A)** Dysmenorrhea Symptom Score: presented as mean ± SD at baseline and after Cycles 1–3 for the EGT (blue) and Placebo (orange) groups. *p < 0.05, ***p < 0.001 vs. baseline; †p = 0.028 vs. placebo at Cycle 3. (**B**) COX Menstrual Symptom Scale (CMSS) Score: presented as mean ± SD. EGT significantly reduced total CMSS scores relative to baseline from Cycle 1 onward.

### 3.4 Exploratory Outcomes 3.4.1 Serum Inflammatory Biomarkers

Serum concentrations of IL-6, TNF-α, IL-1β, and PGF2α were measured at baseline (V0) and after cycle 3 (V3). In the placebo group, IL-6 showed a statistically significant decrease (p = 0.0064), while all other biomarkers did not change significantly. In the EGT group, none of the four biomarkers exhibited significant within-group change. The change (V0 − V3) in each biomarker did not differ significantly between the EGT and placebo groups. Spearman’s correlation analysis revealed no significant association between the change in any inflammatory biomarker and the reduction in VAS score. These results suggest that EGT’s analgesic benefit operates independently of modulation of the classical IL-6/TNF-α/IL-1β/PGF2α inflammatory axis, possibly reflecting broader antioxidant and cytoprotective mechanisms. (Figures S1–S3)

### 3.5 Safety and Tolerability

Oral administration of EGT capsules at 120 mg/day exhibited an excellent safety and tolerability profile. No adverse events (AEs) or serious adverse events (SAEs) were reported in either group. Vital sign measurements and laboratory parameters showed no clinically meaningful or statistically significant changes within either group (Figure S5).

### 3.6 Responder Analysis

A clinically meaningful responder was defined as a patient achieving ≥30% or ≥50% reduction from baseline VAS score at cycle 3, consistent with IMMPACT consensus recommendations and foundational methodologies for pain trials [13, 14]. Using the ≥30% threshold, 84.2% of EGT-treated patients (16/19) were classified as responders versus 40.0% in the placebo group (Fisher’s exact test, p = 0.008). With the ≥50% threshold, the EGT responder rate remained 84.2%, compared with 35.0% for placebo (p = 0.003). The number needed to treat (NNT) for ≥50% VAS reduction was 2.0 (95% CI: 1.4–3.7), indicating a clinically relevant intervention effect. (Figure 4)

**Figure 4.**
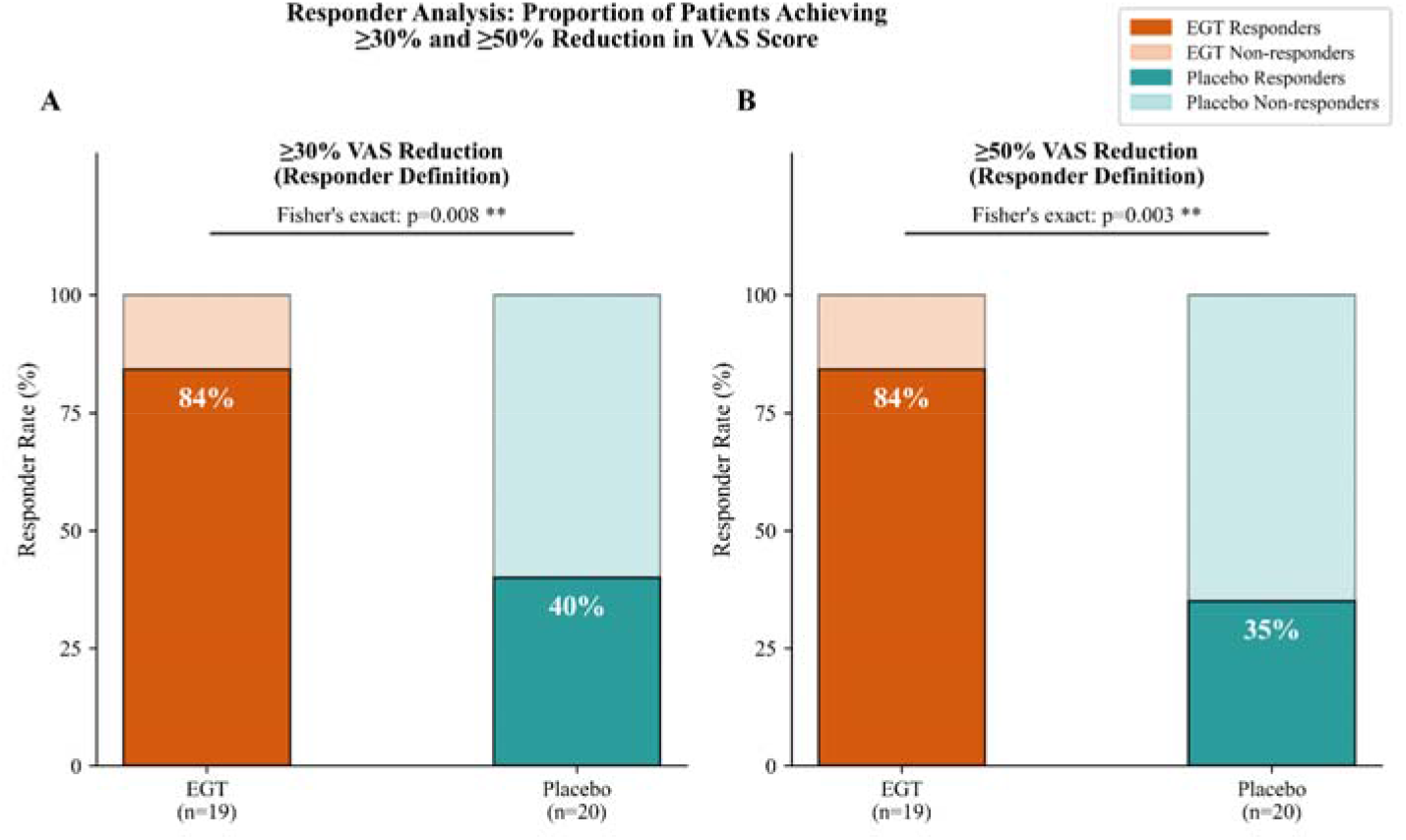
Responder analysis for pain reduction. The proportion of patients in the EGT (n = 19) and Placebo (n = 20) groups achieving a ≥30% (left panel) and ≥50% (right panel) VAS reduction from baseline to Cycle 3. Dark and light bars represent responders and non-responders, respectively. P-values were calculated using Fisher’s exact test. The number needed to treat (NNT) for a ≥50% VAS reduction is 2.0 (95% CI: 1.4–3.7).

### 3.7 Longitudinal Time-Trend Analysis (Linear Mixed Model)

A linear mixed model (LMM) was fitted to test for differential treatment effects over time. The Time × Group interaction was highly statistically significant (β = −0.587, p < 0.001; Figure 5), indicating that the EGT group declined at a significantly steeper rate per cycle than the placebo group. The group main effect at baseline did not reach statistical significance (p = 0.053), confirming comparability at V0. These findings support the conclusion that EGT produced an accelerating analgesic benefit with each successive cycle. (Figure 5)

**Figure 5.**
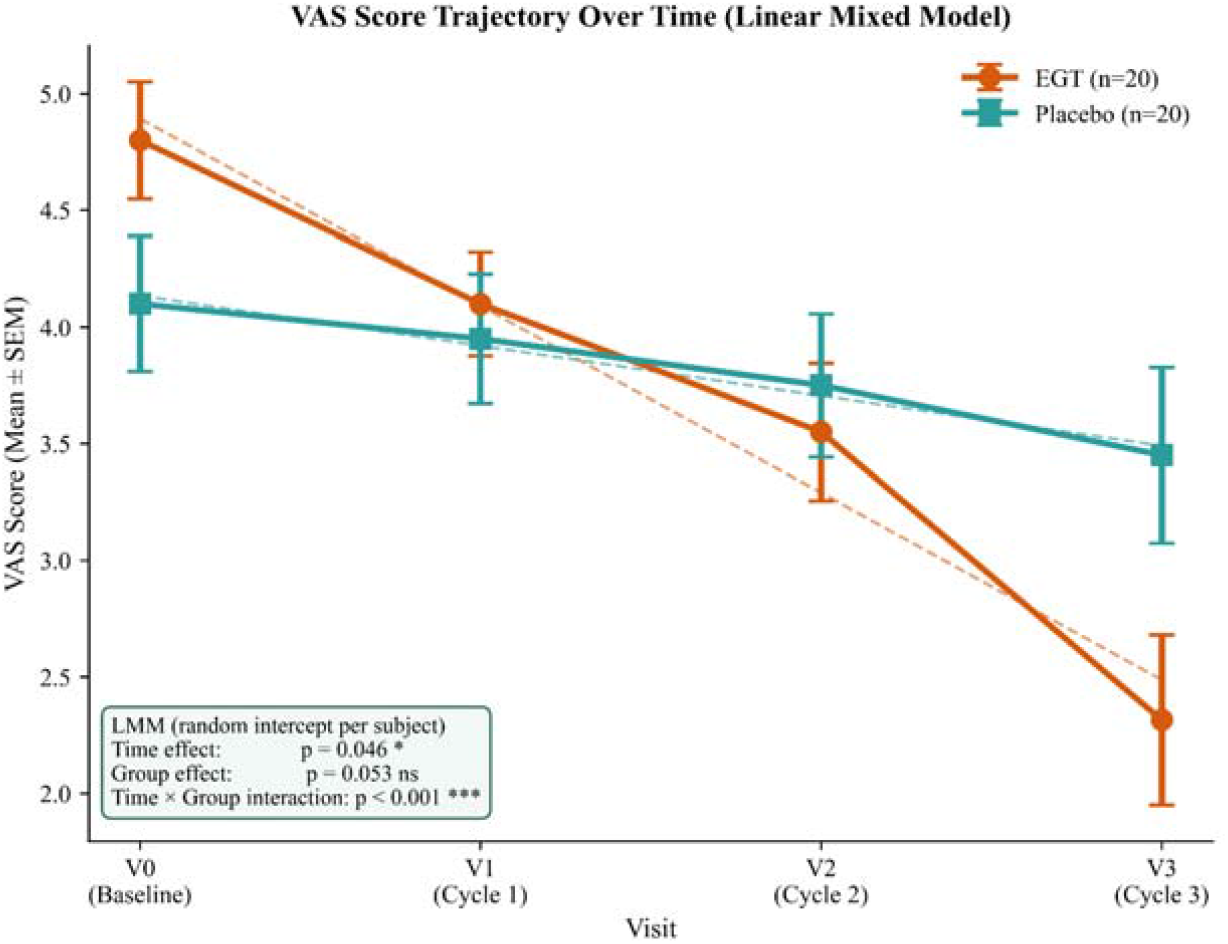
VAS score trajectories over four visits (V0– V3) modelled by a linear mixed model (LMM). Points and error bars represent the observed group mean ± standard error of the mean (SEM) (EGT: blue circles; Placebo: orange squares). Dashed lines illustrate LMM-fitted trajectories. The inset box reports the LMM fixed-effect p-values, with the Time × Group interaction showing a highly significant difference (***p < 0.001),confirming a significantly faster rate of pain reduction in the EGT group relative to the placebo group.

### 3.8 Covariate and Subgroup Analysis

Spearman’s rank correlation between baseline VAS score and absolute VAS reduction was significant in both groups, indicating that patients with higher baseline pain tended to show greater absolute improvement regardless of group allocation, consistent with regression to the mean. Within the EGT group, participants dichotomized into high-baseline (VAS ≥ 5) and low-baseline (VAS < 5) subgroups showed no statistically significant difference in absolute VAS reduction (p = 0.204), suggesting EGT provides consistent benefit across varying baseline pain severities. (Figure S4)

## 4. Discussion

To our knowledge, this is the first randomized, double-blind, placebo-controlled trial to evaluate the clinical efficacy and safety of oral L-ergothioneine (EGT) in women with primary dysmenorrhea. Following a 3-consecutive-cycle intervention, EGT supplementation (120 mg/day) significantly reduced both peak menstrual pain (VAS) and systemic symptom burdens compared to baseline and placebo. The clinical benefits of EGT appeared cumulative, strengthening progressively from the first to the third cycle.

The absence of significant between-group differences in serum IL-6, TNF-α, IL-1β, and PGF2α suggests EGT’s analgesic mechanism may not operate primarily by suppressing these specific circulating markers. This is biologically plausible: EGT concentrates intracellularly via the OCTN1 transporter, quenching reactive oxygen species and stabilizing mitochondrial function at the subcellular level. Previous studies emphasize that oxidative stress markers, rather than classic cytokines, are more reliably perturbed in PD [15]. Thus, EGT’s primary benefit is likely mediated through antioxidant pathways upstream of prostaglandin biosynthesis. Future studies should incorporate measures of oxidative stress and endometrial prostaglandin levels to clarify this mechanism.

In the broader landscape of nutritional interventions for primary dysmenorrhea, conventional dietary strategies incorporating micronutrients such as Vitamin E, Omega-3 fatty acids, or zinc have been widely explored [8]. However, EGT presents a distinct mechanistic advantage over these traditional nutrients. While classical dietary antioxidants often struggle to achieve efficient intracellular concentrations at the specific sites of acute oxidative damage, EGT is actively transported and concentrated directly into mitochondria via the OCTN1 transporter [10]. This highly targeted subcellular accumulation allows EGT to quench reactive oxygen species at their primary source of generation, potentially explaining the robust and progressive clinical analgesic benefits observed in our trial.

During menstruation, declining progesterone triggers arachidonic acid release and prostaglandin synthesis, generating localized oxidative stress. Elevated ROS perpetuates a cycle of inflammation, tissue hypoxia, and pain sensitization. By scavenging free radicals via OCTN1-mediated mitochondrial accumulation, EGT likely interrupts this positive feedback loop, promoting myometrial relaxation and ameliorating ischemic pain.

Furthermore, the excellent safety profile observed aligns with EGT’s status as a safe dietary derivative, offering a gentler stabilization of the uterine environment without the systemic risks associated with conventional NSAIDs.

### Limitations

Limitations include the relatively small single-center sample size (N = 40) and the brief 3-month follow-up, which precludes assessment of long-term benefits or relapse rates. Additionally, exploratory objective biomarkers collected will be subjected to dedicated analysis in a subsequent independent manuscript. Future large-scale, multicenter trials are warranted to validate these findings.

## 5. Conclusion

Oral supplementation with 120 mg/day of L-ergothioneine over three consecutive menstrual cycles significantly and progressively alleviates peak menstrual pain and associated systemic symptoms in women with primary dysmenorrhea. EGT emerges as a highly promising, well-tolerated, non-pharmacological nutritional intervention. Future large-scale trials will further elucidate the full nutritional potential of EGT in women’s reproductive health.

## Ethics Approval and Consent to Participate

This study was conducted in strict adherence to the Declaration of Helsinki and Good Clinical Practice (GCP) guidelines. The study protocol (Protocol No. MJLYJN-IIT-25009) was approved by the Clinical Trial Ethics Committee of Qingdao Central Hospital of Rehabilitation University. Written informed consent was obtained from all participants prior to screening and enrollment.

## Clinical Trial Registration

This trial was registered with the Chinese Clinical Trial Registry (Registration Number: ChiCTR2500112557) on November 17, 2025 (Retrospectively registered).

## Data Availability Statement

The datasets generated and/or analyzed during the current study are not publicly available due to participant privacy but are available from the corresponding author upon reasonable request.

## Authors’ Contributions

G.X., W.D., and F.L. conceptualized and designed the study. C.G., and T.T. conducted the clinical trial and collected the data. J.C., W.L., and T.T. analyzed the data and performed the statistical analysis. W.L. wrote the first draft of the manuscript. All authors reviewed, edited, and approved the final manuscript.

