## Supplementary_Materials for "Efficacy and Safety of Oral Gene III^®^ L-Ergothioneine Capsules in Primary Dysmenorrhea: A Randomized, Double-Blind, Placebo-Controlled Clinical Trial" (from another version of this preprint: version 1, 2026-03-27)

**Figure S1**

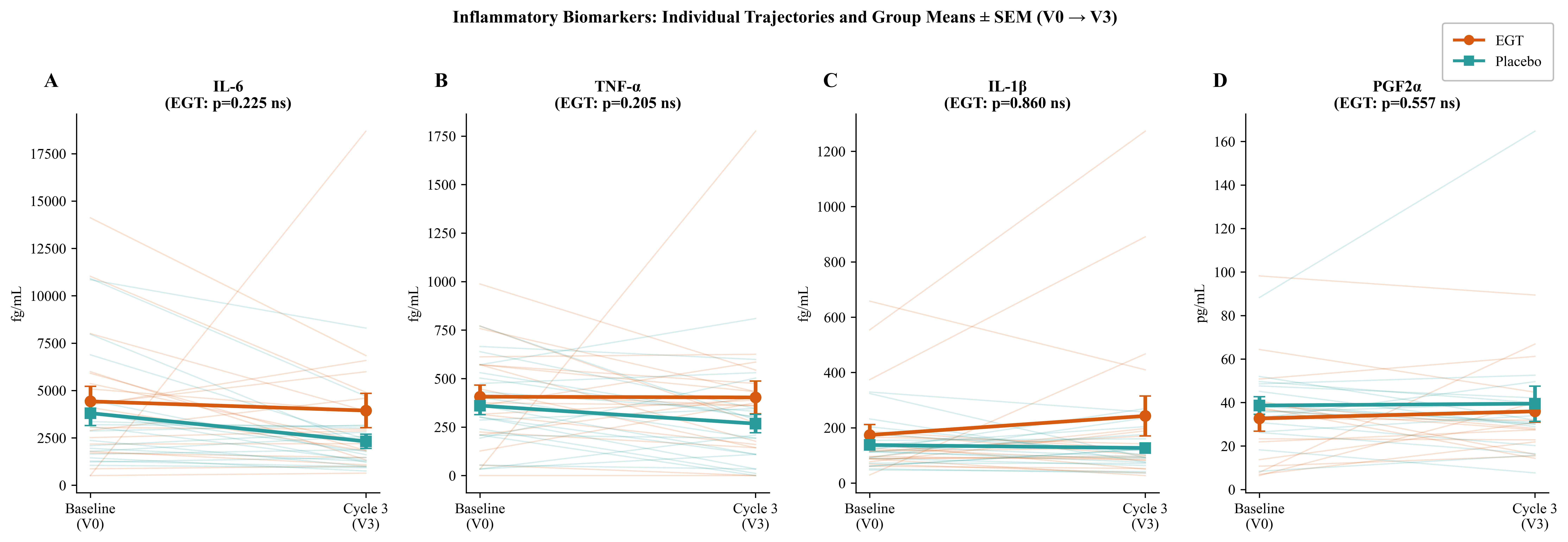

**Figure S1. Serum inflammatory biomarker trajectories from baseline (V0) to Cycle 3 (V3).**

Light grey/blue lines represent individual patient trajectories. Bold lines with error bars indicate group mean ± SEM (EGT: orange circles, n=19; Placebo: teal squares, n=20). Within-group p-values from the Wilcoxon signed-rank test (or paired t-test) are shown above each panel. None of the four biomarkers showed a statistically significant within-group change in the EGT group (all p≥0.205). In the placebo group, IL-6 exhibited a significant within-group decrease (p = 0.006). Abbreviations: IL-6, interleukin-6; TNF-α, tumor necrosis factor-α; IL-1β, interleukin-1β; PGF2α, prostaglandin F2α.

**Figure S2**

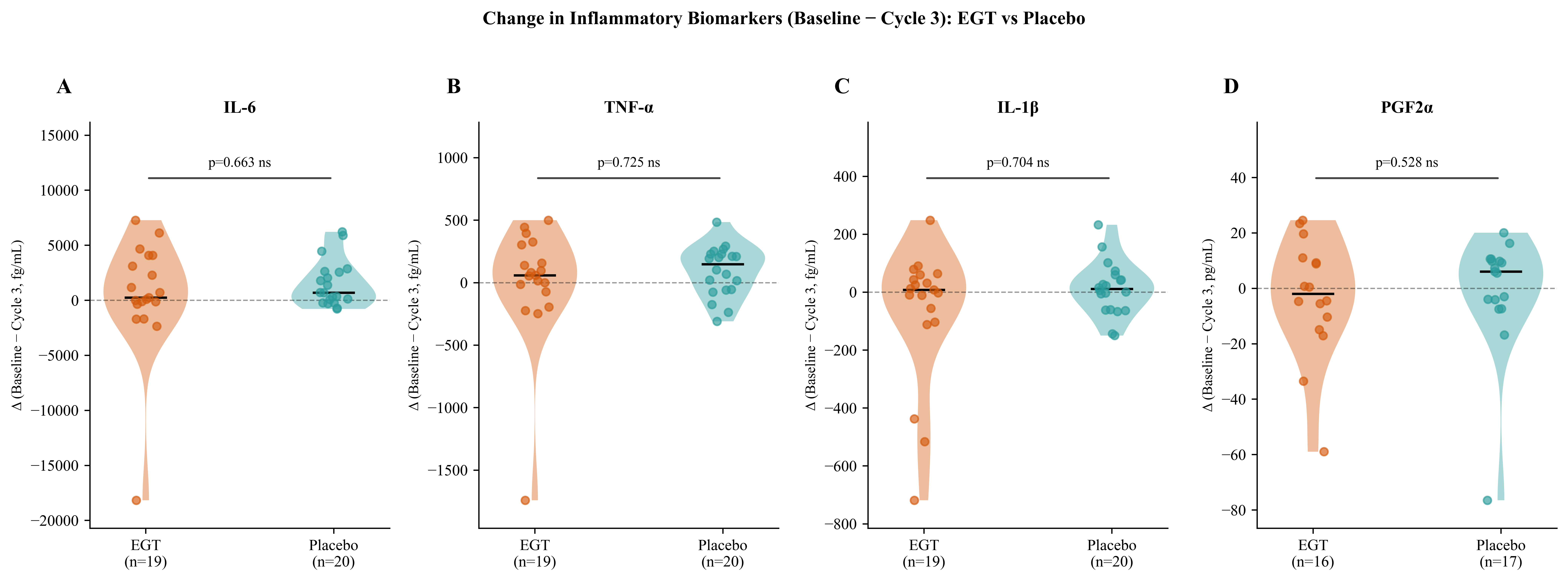

Figure S2. Figure S2. Distribution of inflammatory biomarker changes (Δ=Baseline − Cycle 3) in the EGT (orange) and Placebo (teal) groups, displayed as violin plots overlaid with individual data points.

Horizontal bars indicate group medians. The dashed line at zero indicates no change. Between-group comparisons were performed by Mann–Whitney U test; no significant between-group difference was detected for any biomarker (IL-6: p = 0.663; TNF-α: p = 0.725; IL-1β: p = 0.704; PGF2α: p = 0.528).

**Figure S3**

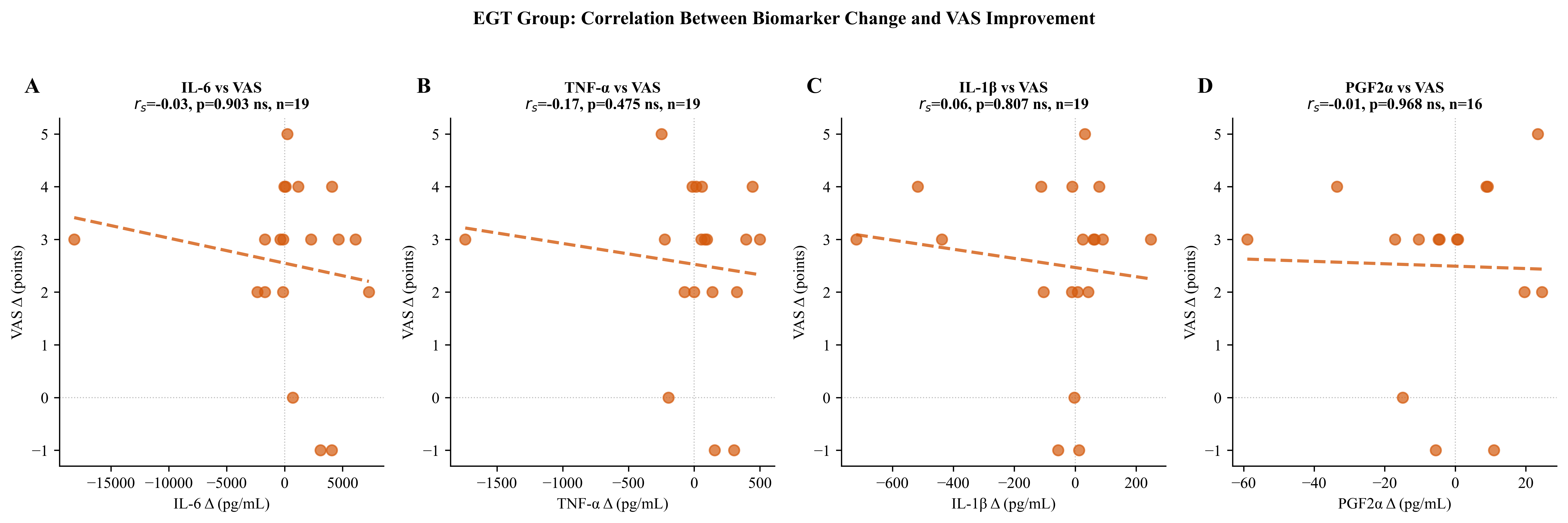

Figure S3. Figure S3. Spearman’s rank correlation between the change in each serum inflammatory biomarker (Δ = V0 − Cycle 3, pg/mL) and VAS improvement (Δ VAS = V0 − Cycle 3, points) in the EGT group.

Individual data points are shown; dashed lines represent ordinary least-squares regression fits. Correlation coefficients (ρ_s_) and p-values are displayed in each panel title. None of the four biomarkers correlated significantly with VAS reduction (IL-6: ρ_s_ = -0.03, p = 0.903; TNF-α: ρ_s_ = -0.17, p = 0.475; IL-1β: ρ_s_ = 0.06, p = 0.807; PGF2α: ρ_s_ = -0.01, p = 0.968, n=16), suggesting that EGT’s analgesic benefit operates independently of these classical inflammatory mediators.

**Figure S4**

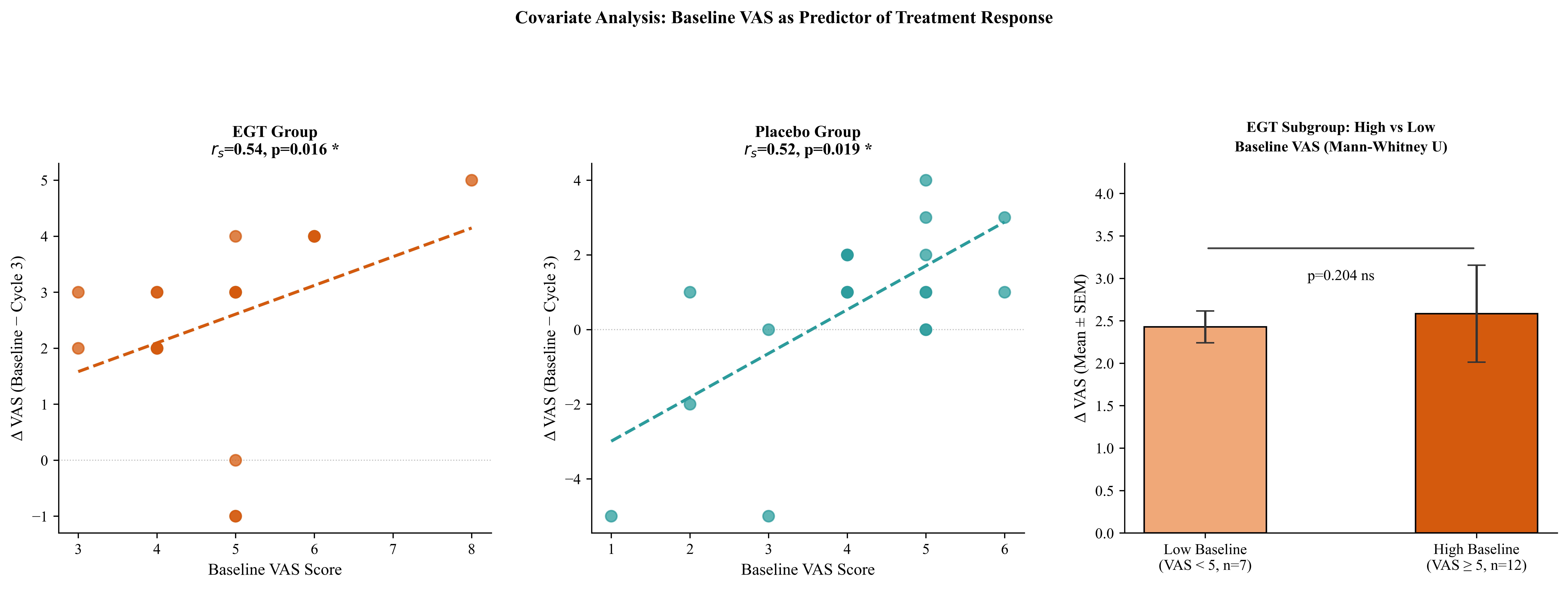

Figure S4. Covariate and subgroup analysis.

(A) Spearman’s correlation between baseline VAS score and absolute VAS reduction (V0 − Cycle 3) in the EGT group (ρ_s_ = 0.544, p = 0.016). (B) Corresponding correlation in the Placebo group (ρ_s_ = 0.520, p = 0.019). In both groups, higher baseline pain was associated with greater absolute improvement, consistent with regression to the mean. Dashed lines represent OLS regression fits. (C) Subgroup analysis within the EGT group: mean ± SEM absolute VAS reduction for participants with high baseline VAS (≥5, n = 12) versus low baseline VAS (< 5, n = 7). No statistically significant difference was observed between subgroups (Mann–Whitney U, p = 0.204), suggesting EGT provides consistent benefit across a range of baseline pain severities.

**Figure S5**

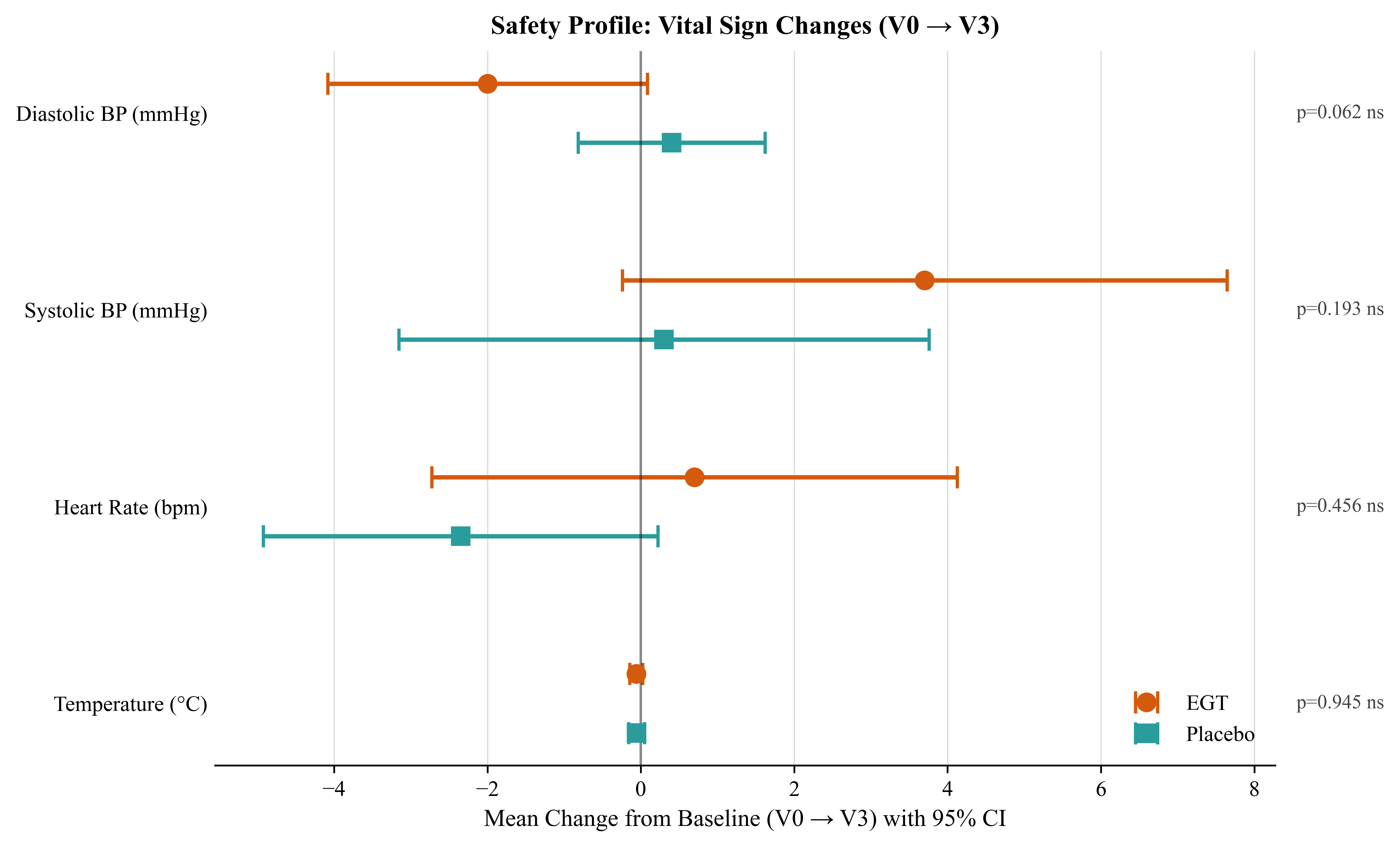

Figure S5. Safety profile: mean change from baseline (V0 → Cycle 3) in vital sign parameters for the EGT group (orange circles, n = 19) and Placebo group (teal squares, n = 20), presented as a forest plot with 95% confidence intervals. Between-group p-values from the Mann–Whitney U test are shown to the right of each parameter. No statistically significant between-group differences were detected for body temperature (p = 0.945), heart rate (p = 0.456), systolic blood pressure (p = 0.193), or diastolic blood pressure (p = 0.062). These data confirm that oral EGT at 120 mg/day did not produce clinically meaningful cardiovascular or thermoregulatory changes over three consecutive menstrual cycles. SBP, systolic blood pressure; DBP, diastolic blood pressure.
